# Preliminary estimation of the novel coronavirus disease (COVID-19) cases in Iran: a modelling analysis based on overseas cases and air travel data

**DOI:** 10.1101/2020.03.02.20030320

**Authors:** Zian Zhuang, Shi Zhao, Qianying Lin, Peihua Cao, Yijun Lou, Lin Yang, Daihai He

## Abstract

As of 1 March 2020, Iran has reported 987 COVID-19 cases and including 54 associated deaths. At least six neighboring countries (Bahrain, Iraq, Kuwait, Oman, Afghanistan and Pakistan) have reported imported COVID-19 cases from Iran. We used air travel data and the cases from Iran to other Middle East countries and estimated 16533 (95% CI: 5925, 35538) COVID-19 cases in Iran by 25 February, before UAE and other Gulf Cooperation Council countries suspended inbound and outbound flights from Iran.

## Introduction

The coronavirus disease 2019 (COVID-19) first emerged in Wuhan, Hubei Province, China in the end of 2019, and soon spread to the rest of China and overseas (Bogoch *et al*., 2020). To date, there were 78630 cases reported in China and 3664 cases confirmed in 46 foreign countries (World Health Organization, 2020). Recently, Iran has become an epicenter in the Middle East region, which has the largest cumulative number of deaths outside China (Wikipedia, 2020). As of 1 March 2020, Iran has confirmed 987 COVID-19 cases and including 54 associated deaths (Wikipedia, 2020). At least six neighboring countries (Bahrain, Iraq, Kuwait, Oman, Afghanistan and Pakistan) have reported imported COVID-19 cases from Iran. Given the serious concerns over the under-ascertainment of COVID-19 cases in Iran (ABC News, 2020), UAE and other Gulf Cooperation Council countries have suspended inbound and outbound flights from Iran since 25 February 2020 (Khaleej Times, 2020).

## Objective

In this study, we used the imported cases and air travel data from Iran to other Middle East countries to estimate the number of COVID-19 cases in Iran. Then we compared our estimates with the number of reported cases in Iran to evaluate the extent of under-ascertainment.

## Data

We obtained transport capability of international airlines from 30 major airports in Iran from 1 February to 24 February 2020 (WorldData, 2020), from the Variflight platform (https://data.variflight.com/). We collected the number of exported cases from Iran to other countries in the Middle East (World Health Organization, 2020). Population size of Iran (81,800,269 in 2018) was obtained from the World Bank (https://data.worldbank.org/). Table 1 shows the number of daily population flow and total reported cases of countries used in our calculation.

**Table 1:**
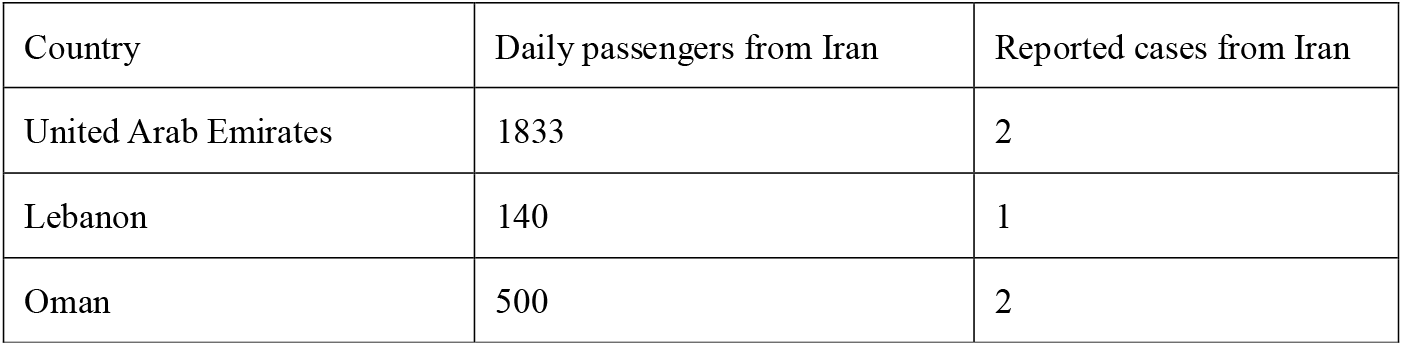
Number of daily population flow and total reported cases of countries used in the estimation, 25 February 2020

| Country | Daily passengers from Iran | Reported cases from Iran |
| --- | --- | --- |
| United Arab Emirates | 1833 | 2 |
| Lebanon | 140 | 1 |
| Oman | 500 | 2 |

## Methods

Following Imai *et al*.’s (2020), we assumed number of cases (*n*) exported from Iran follows a Binomial distribution (Bin) with size *N* and probability *p*. Here, *N* represents total number of cases infected in Iran, and *p* is the probability that one case is detected overseas. We approximated *p* by the ratio of daily outbound passengers of Iran (*D*) over total size of population (*M*) that those airports serve and multiplying the mean duration (*t*) from exposure to detection. Thus, *p* = *Dt* / *M*. We set *t* at 10 days according to (Imai *et al*., 2020). We assumed that the catchment population is the total population of Iran. We calculated the maximum likelihood estimates of the total number of COVID-19 cases in Iran (λ) by fitting model to the number of confirmed cases with Binomial-distributed likelihood framework. As shown in Eqn (1), *l*(·) represents the total log-likelihood and k is the total number of countries we selected. The 95% confidence intervals (CI) are calculated by using the profile likelihood estimation approach determined by a Chi-square quantile.

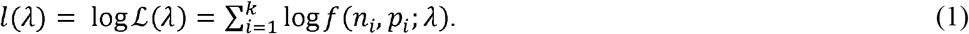

To explore more alternative scenarios, we also considered a smaller catchment population (60,000,000) and a shorter detection window (8 days). We considered a baseline scenario with 100% attendance rate for each aircraft as well as other alternative scenarios. In addition, we tested the situation that 90% and 70% attendance rate for each aircraft, keeping other situation as baseline scenario.

## Results and discussion

We estimated the total number of cases in Iran was 16533 (95% CI: 5925, 35538) by 25 February 2020. Table 2 summarizes the sensitivity analysis of varying the baseline assumptions and alternative scenarios about catchment population, detection window and attendance rates.

**Table 2:** Estimated case numbers based on the baseline assumptions and alternative scenarios.

|  | Baseline | Smaller catchment | Shorter detection window | 90% load factors | 70% load factors |
| --- | --- | --- | --- | --- | --- |
| Effective catchment population | 81800269 | 60000000 | 81800269 | 81800269 | 81800269 |
| Detection window (day) | 10 | 10 | 8 | 10 | 10 |
| total cases | 16533 (5925, 35538) | 12125 (4345, 12145) | 20667 (7408, 44424) | 18368 (6583, 39482) | 23627 (8472, 50780) |

As suggested by Zhao *et al*. (2020), underreporting was likely occurred at the early stage of the COVID-19 outbreak, due to the shortage of diagnostic kits and inadequate screening for suspected cases (Imai *et al*., 2020). Recent study by Tuite *et al*. (2020) estimated a total of 18,300 (95% CI: 3770-53,470) cases in Iran by 25 February 2020. Our estimates are consistent with Tuite *et al*.’s (2020) work. To conclude, Iran’s ascertainment rate could be at a level of 0.6% by Feb 25, 2020. The health security capabilities of many countries in the Middle East are below the world average (Ghsindex, 2019), which means that if Iran’s epidemic continues to spread to neighboring countries, the fragile public health systems of Middle East countries is likely to find difficulty in coping with the outbreak.

## Data Availability

We used only publicly available data.

## Declarations

### Ethics approval and consent to participate

The ethical approval or individual consent was not applicable.

### Availability of data and materials

All data and materials used in this work were publicly available.

### Consent for publication

Not applicable.

## Acknowledgements

None.

## Disclaimer

The funding agencies had no role in the design and conduct of the study; collection, management, analysis, and interpretation of the data; preparation, review, or approval of the manuscript; or decision to submit the manuscript for publication.

